# Auxiliary Clinical Prompt Integration into Vision-Language Prompt SAM for Brain Tumor Segmentation

**DOI:** 10.64898/2026.04.15.26351001

**Authors:** Yuto Hakata, Miko Oikawa, Shin Fujisawa

## Abstract

**Background:** Adult diffuse glioma is a representative class of primary brain tumors for which accurate MRI-based tumor segmentation is indispensable for treatment planning. Conventional automated segmentation methods have relied primarily on image information and spatial prompts, and auxiliary clinical information that is routinely acquired in clinical practice has not been sufficiently exploited as an input.

**Objective:** Building on a **dual-prompt-driven Segment Anything Model (SAM) extension framework [20]** that fuses visual and language reference prompts, we propose a method that integrates patient demographics, unsupervised molecular cluster variables derived from TCGA high-throughput profiling, and histopathological parameters as learnable prompt embeddings, and we evaluate its effect on the accuracy of lower-grade glioma (LGG) MRI segmentation.

**Methods:** An auxiliary prompt encoder converts clinical metadata into high-dimensional embeddings that are fused with the prompt representations of Segment Anything Model (SAM) ViT-B through a cross-attention fusion mechanism. The TCGA-LGG MRI Segmentation dataset (Kaggle release by Buda et al. [24]; n = 110 patients; WHO grade II–III) was split at the patient level (train/val/test = 71/17/22) using three different random seeds, and the three slices with the largest tumor area were extracted from each patient. To avoid pseudo-replication arising from multiple slices per patient and repeated measurements across seeds, **our primary analysis aggregated Dice and 95th-percentile Hausdorff distance (HD95) to the patient × seed unit (n = 66)**; secondary analyses at the unique-patient level (n = 22) and at the per-slice level (n = 198) are also reported. Pairwise comparisons used **paired t-tests** with Bonferroni correction (k = 3) and **Wilcoxon signed-rank tests**, and a **permutation test (K = 30)** served as an auxiliary check of effective use of the auxiliary information.

**Results:** At the patient × seed level (n = 66), Proposed (full clinical) achieved a Dice gain of Δ = +0.287 over the zero-shot SAM ViT-B baseline (paired-t p = 4.2 × 10⁻¹⁵, Cohen’s d_z = +1.25, Bonferroni-corrected p ≪ 0.001; Wilcoxon p = 2.0 × 10⁻¹⁰), and HD95 improved from 218.2 to 64.6. Because zero-shot SAM is not designed for domain-specific medical segmentation, the large absolute HD95 gap largely reflects the expected domain gap rather than a competitive baseline. The additional contribution of the full clinical configuration over the demographics-only configuration was Δ Dice = +0.023 (paired-t p = 0.057, Bonferroni-corrected p = 0.172), which **did not reach statistical significance** at the patient level and is reported as a directional trend. The permutation test (K = 30, seed 2025) yielded real-metadata Dice = 0.819 versus a shuffled-metadata mean of 0.773, giving an empirical p = 0.032 = 1/(K + 1), which is at the resolution limit of this test and should therefore be interpreted as preliminary evidence.

**Conclusions:** Integrating auxiliary clinical information as multimodal prompts produced a large improvement over the zero-shot SAM baseline on this LGG cohort. More importantly, a robustness analysis showed that Proposed (full clinical) **outperformed the trained Base (no auxiliary information) under all tested spatial-prompt conditions**, including perfect centroid (Δ = +0.014), and that the advantage was most pronounced in the prompt-free regime (Δ = +0.231, p = 0.039), where the base model collapsed but the proposed model maintained meaningful segmentation by leveraging clinical metadata alone. The additional contribution of molecular and histopathological information beyond demographics was not statistically resolved at the patient level (Δ = +0.023, n.s.). Establishing clinical utility will require external validation on larger multi-center cohorts and direct comparisons with established segmentation methods.

## 1. Introduction

Adult diffuse glioma is a class of primary brain tumors that, under the 2021 WHO Classification of Tumors of the Central Nervous System [3], is now reorganized on the basis of molecular markers such as IDH mutation status and 1p/19q co-deletion. MRI-based tumor segmentation plays a central role in planning treatment for these tumors [1, 4]: accurate delineation of tumor boundaries is essential for determining the extent of surgical resection, defining target volumes in radiotherapy, and performing longitudinal volumetric assessment of treatment response [4, 5]. In particular, for glioblastoma (glioblastoma, IDH-wildtype; WHO CNS grade 4) the median survival under standard temozolomide-based chemoradiotherapy remains approximately 14.6 months [2], and precise treatment planning directly impacts prognosis. The experimental validation of this work, however, is limited to WHO grade II–III lower-grade gliomas (LGG), and the findings reported below should be interpreted within that scope.

Traditionally, this segmentation has relied on manual annotation by experienced neuroradiologists. However, manual annotation imposes a substantial burden in terms of both time and effort, with inter-rater disagreement rates reaching 20–30% in complex cases [5, 6]. The global shortage of neuroradiologists further exacerbates this problem, creating a strong demand for robust automated or semi-automated segmentation techniques [7].

Advances in deep learning have addressed this need. Encoder–decoder architectures pioneered by U-Net [8], along with Vision Transformer–based models exemplified by TransUNet [12] and Swin-UNet [13], have been successively proposed, contributing to improvements in brain tumor segmentation accuracy. In 2023, Meta AI Research released the Segment Anything Model (SAM) [15], which was trained on large-scale natural image data and opened up new possibilities for prompt-based general-purpose segmentation. However, directly applying SAM to medical images has been reported to yield degraded performance on low-contrast boundaries and complex tumor morphologies [16, 17], and medical-domain adaptations such as MedSAM [18] and SAM-Med2D [19] have accordingly been developed.

Most of these methods use only images and spatial prompts as inputs to the segmentation model, and the input information is deliberately curated. This curation is motivated by practical considerations such as avoiding the introduction of noise and keeping the model design simple. In actual clinical practice, however, physicians interpret imaging findings by integrating them with multidimensional information about the patient, including demographics, IDH mutation status, and MGMT promoter methylation status [7]. These clinical factors have been reported to correlate with the imaging phenotypes of tumors [21, 22] and can serve as auxiliary discriminative cues that are not obtainable from images alone. A strength of deep learning, and of attention-based models in particular, lies in their ability to learn and exploit latent associations among seemingly unrelated heterogeneous information sources. Nevertheless, such structured clinical information has rarely been used as input to segmentation models.

Motivated by this observation, we focused on a dual-prompt-driven SAM extension framework that fuses visual and language reference prompts (Vision-Language Prompt SAM, VLP-SAM) [20]. This framework has demonstrated the effectiveness of multimodal prompting on standard benchmarks, and we hypothesized that extending it by incorporating auxiliary clinical information as additional prompt embeddings would further improve the accuracy of LGG MRI segmentation. Specifically, we designed a method that integrates patient demographic data, unsupervised molecular cluster variables, and histopathological classification parameters as learnable prompt embeddings, and we systematically evaluated the effect of adding auxiliary information on segmentation accuracy in experiments on the publicly available TCGA-LGG brain tumor dataset.

## 2. Methods

### 2.1 Study design and reporting

This is a retrospective methodological investigation based on secondary analysis of a publicly available, anonymized medical imaging dataset. It does not involve recruitment of new patients, any intervention, or handling of identifiable personal data. The manuscript is reported in accordance with the CLAIM (Checklist for Artificial Intelligence in Medical Imaging) guidelines.

### 2.2 Dataset

We used the TCGA-LGG MRI Segmentation dataset released on Kaggle by Buda et al. [24] (n = 110 patients; WHO grade II–III), which is part of the TCGA brain tumor imaging collection made publicly available through The Cancer Imaging Archive (TCIA) [23]. For each patient, FLAIR sequence slices and the corresponding tumor masks were provided. Regarding the provenance of the ground-truth segmentation masks, we used the expert annotations supplied with the Kaggle release; these are understood by the authors to follow the two-stage expert annotation process developed for TCGA glioma collections as described by Bakas et al. [24].

The dataset was split into training, validation, and test sets at the patient level in a 71/17/22 ratio, with stratified sampling by histopathological grade. Experiments were repeated with three different random seeds (13, 42, 2025), and the three slices with the largest tumor area were extracted from each patient. Consequently, the test set for each seed contained 22 patients × 3 slices = 66 slice samples, and pooling across the three seeds yielded **a total of 198 slice observations (with a unique patient count of 56 across all test folds)**. The patient-level split completely precludes data leakage between the training, validation, and test sets.

### 2.3 Auxiliary clinical information

A distinctive feature of the proposed method is that it incorporates auxiliary clinical information, extracted from the TCGA metadata repository, as supplementary prompt input to the segmentation model. The auxiliary information used is grouped into the following three categories.

- **Demographics:** age at diagnosis (age_at_initial_pathologic) and biological sex (gender).
- **Molecular cluster variables:** MethylationCluster, RNASeqCluster, and miRNACluster. These variables are all **unsupervised consensus cluster assignments** derived by the TCGA Research Network from high-throughput molecular profiling (genome-wide DNA methylation, RNA-Seq, and miRNA-Seq) [25] and should be **distinguished from the direct molecular markers** (IDH mutation, 1p/19q co-deletion, MGMT promoter methylation by direct assay) specified by the 2021 WHO Classification of Tumors of the Central Nervous System [3]. In particular, MethylationCluster reflects genome-wide DNA methylation patterns based on thousands of CpG sites and is not a surrogate indicator specific to MGMT promoter methylation alone. We adopted these cluster variables as additional auxiliary cues because prior TCGA reports [25] indicate that they are related to, while distinct from, the molecular subtypes specified by the 2021 WHO classification.
- **Histopathology:** WHO grade (neoplasm_histologic_grade), histological type (histological_type), tumor location (tumor_location), and laterality (laterality) [25].

All of these variables are routinely acquired in clinical practice, and we adopted them on the grounds that they can provide cues about tumor characteristics that are not obtainable from images alone. In this study, we used only actual values contained in the public dataset, and missing values were handled as a dedicated “unknown” index; no pseudo data were generated.

### 2.4 Model architecture

The proposed method adopts the vision-language prompt-driven SAM extension framework (VLP-SAM) [20] as its base architecture. This base architecture consists of SAM’s image encoder (a ViT-B backbone in our implementation), a dual-modal prompt encoder, and a mask decoder, and is characterized by a cross-attention layer that fuses visual prompts with auxiliary prompt representations.

The central contribution of this study is the design and integration of an **auxiliary prompt encoder** that converts clinical metadata into high-dimensional prompt embeddings. This module consists of parallel encoding paths. Continuous variables such as age are processed with sinusoidal positional encoding (dim = 64), whereas categorical variables such as sex and clustering information are processed with trainable embedding layers (nn.Embedding, 32-d each). The outputs of the individual paths are concatenated and then projected into a shared prompt embedding space (d_model = 256, num_tokens = 4) by a two-layer MLP with GELU activation.

The resulting auxiliary prompt embeddings are integrated with the sparse prompt representations of the base architecture through a **cross-attention fusion mechanism**. Specifically, multi-head cross-attention (8 heads, dropout = 0.1) is applied with the sparse_embeddings of the SAM prompt encoder as queries and the auxiliary prompt embeddings as keys and values, followed by a residual connection, layer normalization, and a feed-forward network, yielding the final fused prompt representation. This design is based on the principle that the auxiliary information should modulate rather than replace the spatial prompt representation.

The mask decoder receives the fused image embeddings and the integrated prompt embeddings and produces a segmentation mask through transformer decoder layers and spatial upsampling.

### 2.5 Training protocol

We trained the model using a composite loss function combining Dice loss and binary cross-entropy loss. The SAM ViT-B image encoder weights were frozen, and the auxiliary prompt encoder, cross-attention fusion layer, SAM prompt encoder, and mask decoder were trained end-to-end for 40 epochs using the AdamW optimizer (lr = 1 × 10⁻⁴, weight_decay = 1 × 10⁻⁴) with a cosine annealing learning-rate schedule. Training was performed independently with three different random seeds (13, 42, 2025). All experiments were carried out on a single NVIDIA RTX 4060 Laptop GPU (8 GB VRAM). With the image encoder features pre-cached, inference (prompt encoding + cross-attention fusion + mask decoding) required approximately 0.02 seconds per slice; the full pipeline including the image encoder forward pass required approximately 0.2 seconds per slice on the same hardware.

### 2.6 Evaluation metrics and statistical analysis

We compared the following three configurations:

- (i) untrained **SAM ViT-B zero-shot** as a lower-bound baseline;
- (ii) **Proposed (demographics)**, which uses only demographic information as auxiliary prompts;
- (iii) **Proposed (full clinical)**, which uses all categories of clinical information as auxiliary prompts.

Segmentation performance was quantitatively evaluated using the Dice similarity coefficient (DSC) and the 95th-percentile Hausdorff distance (HD95).

### Unit of statistical analysis

The per-slice observations (3 seeds × 22 patients × 3 slices = 198 observations) are not independent: slices from the same patient, and observations from the same patient under different seeds, are strongly correlated. Applying paired t-tests directly at the per-slice level would therefore overestimate the effective degrees of freedom (pseudo-replication) and produce artificially small p-values. To avoid this, **the primary analysis aggregated Dice and HD95 to the (seed, patient) level by averaging across the three slices, yielding n = 66 patient-seed observations**. Secondary robustness checks were performed at the unique-patient level (n = 22, averaged across seeds as well as slices) and at the per-slice level (n = 198), and these are reported as supplementary evidence.

### Statistical tests

Pairwise comparisons used **paired t-tests** and, as a non-parametric robustness check, **Wilcoxon signed-rank tests**. For the three pairwise comparisons we applied **Bonferroni correction** with k = 3. Effect size was reported as Cohen’s d_z, and 95% confidence intervals were estimated by bootstrap (B = 2,000 percentile intervals). As an additional sensitivity analysis addressing the nested structure of the data (slices within patients, measurements across seeds), we fitted a **linear mixed-effects model (LME)** with a random intercept for patient, using REML estimation via statsmodels (v0.14). This analysis uses the patient-seed-level Dice difference as the dependent variable and tests whether the fixed intercept (i.e., the population-average Dice improvement) is significantly different from zero.

### Permutation test

To assess whether the trained model effectively utilizes the auxiliary metadata, we performed a permutation test in which the auxiliary metadata were randomly swapped between patients within the test set. Specifically, the empirical p-value was computed as (r + 1) / (K + 1), where r is the number of shuffled trials whose mean Dice was at least as large as the real-metadata mean Dice, using K = 30 shuffling trials. By construction this test has a lower-bound resolution of 1 / (K + 1) ≈ 0.032, and K = 30 was chosen under computational-cost constraints as a preliminary check; the outcome of this test is therefore interpreted as **preliminary evidence of effective use of auxiliary information, not as a strict significance test**.

The significance threshold was set to α = 0.05. All statistical analyses were implemented in Python 3.11 with SciPy.

## 3. Results

### 3.1 Overall segmentation performance

Table 1 shows the aggregated segmentation performance of the three configurations on the TCGA-LGG test set at the primary analysis level (n = 66, patient-seed).

**Table 1.** Segmentation performance on the TCGA-LGG test set at the primary analysis level (n = 66, patient × seed). Δ values are computed against the zero-shot SAM ViT-B baseline.

| Configuration | Dice (± SD) | HD95 (px) | $\Delta$ vs zero-shot | $\Delta$ vs Base | Notes |
| --- | --- | --- | --- | --- | --- |
| Zero-shot SAM ViT-B | 0.538 (0.472–0.601) | 218.2 | — | — | untrained |
| Base (trained, no aux) | 0.830 ± 0.006 | 52.9 | +0.292 | — (ref) | centroid eval |
| Proposed (demo) | 0.802 (0.756–0.844) | 68.8 | +0.264 | –0.028 | age + sex only |
| **Proposed (full clinical)** | **0.844 ± 0.012** | **50.3** | **+0.306 \*\*\*** | **+0.014** | **full molecular** |

\*\*\* Bonferroni-corrected $p \ll 0.001$ (paired t-test, $k = 3$ ); see Table 2 for full statistics.

Among the compared configurations, zero-shot SAM ViT-B exhibited the lowest performance (Dice 0.538, 95% CI [0.472, 0.601]). The proposed method, **Proposed (full clinical)**, achieved a Dice of 0.825 (95% CI [0.790, 0.856]) and substantially improved over the zero-shot SAM baseline by **Δ Dice = +0.287** (patient-seed level n = 66, paired-t p = 4.2 × 10⁻¹⁵, Cohen’s d_z = +1.25, Bonferroni-corrected p ≪ 0.001, Wilcoxon signed-rank p = 2.0 × 10⁻¹⁰; Table 2). The result was robust at the unique-patient level (n = 22, Δ Dice = +0.280, paired-t p = 1.4 × 10⁻¹², Bonferroni-corrected p < 10⁻¹¹) and under a **linear mixed-effects model** with patient as a random intercept (fixed effect Δ = +0.280, t = 9.15, p = 5.7 × 10⁻²⁰, 95% CI [0.220, 0.340]).

**Table 2.** Pairwise tests at the patient × seed level (n = 66; Bonferroni-corrected, k = 3).

| Comparison | $\Delta \text{ Dice}$ | $d_z$ | paired-$t$ $p$ | Bonf. $p$ | sig. |
| --- | --- | --- | --- | --- | --- |
| Proposed (demo) vs zero-shot | +0.264 | +1.18 | $4.6 \times 10^{-14}$ | $1.4 \times 10^{-13}$ | *** |
| Proposed (full) vs zero-shot | +0.287 | +1.25 | $4.2 \times 10^{-15}$ | $1.3 \times 10^{-14}$ | *** |
| Proposed (full) vs Base (centroid) | +0.014 | — | $p=0.132$ ($n=3$) | — | trend |
| Proposed (full) vs Base (no prompt) | +0.231 | — | $p=0.039$ ($n=3$) | — | * |
| Proposed (full) vs Proposed (demo) | +0.023 | +0.24 | 0.057 | 0.172 | n.s. |

HD95 improved from 218.2 px for zero-shot SAM to 64.6 px for Proposed (full clinical). Because zero-shot SAM is a general-purpose foundation model that has not undergone any domain-specific adaptation for medical imaging, the HD95 of 218.2 px effectively indicates that the baseline model fails to localize the tumor region at all; a large fraction of the absolute HD95 gap therefore reflects the **domain gap between an unadapted foundation model and a medically trained model**. The contribution of our study should be interpreted as the gain that results from integrating auxiliary clinical information on top of a base framework that has been adapted to the medical task, and we discuss this explicitly in §4.1 and in the Limitations (§4.4).

### 3.2 Ablation: adding molecular and histopathological information

We compared Proposed (demographics), which uses only demographic information, with Proposed (full clinical), which uses all categories of clinical information. At the primary analysis level (patient × seed, n = 66), Proposed (full clinical) showed Δ Dice = +0.023 over Proposed (demographics), with paired-t p = 0.057 (Bonferroni-corrected p = 0.172), Cohen’s d_z = +0.24, and Wilcoxon signed-rank p = 0.081. This is a **directional trend but does not reach the α = 0.05 significance threshold** in the current cohort. A linear mixed-effects model with patient as a random intercept yielded a consistent result (fixed effect Δ = +0.022, t = 1.75, p = 0.080, 95% CI [−0.003, 0.046]).

For reference, at the per-slice level (n = 198) the nominal p-value for the same comparison was 0.012 (Bonferroni-corrected 0.036); however, this smaller p-value is influenced by the pseudo-replication described in §2.6 (artificially inflated effective degrees of freedom) and is not the scientifically appropriate analysis unit. We therefore conclude that the current cohort does not provide sufficient statistical power to claim that adding molecular and histopathological information yields a significant improvement beyond demographics, and that re-validation in larger, multi-center cohorts is required. The substantial gain from demographics alone relative to zero-shot SAM (§3.1) stands; the additional contribution of molecular and histopathological information remains inconclusive.

### 3.3 Permutation test as preliminary evidence of auxiliary-information use

To examine whether the trained Proposed (full clinical) model effectively utilizes the auxiliary metadata, we performed a permutation test in which the auxiliary metadata were randomly swapped between patients within the test set. For a representative training trajectory (seed 2025), evaluation with real metadata achieved Dice = 0.819, whereas the mean over 30 shuffled-metadata trials dropped to Dice = 0.773. The empirical p-value computed as (r + 1) / (K + 1) with r = 0 and K = 30 equals 1 / 31 ≈ 0.032 (Table 3, Fig. 4).

**Fig. 1.**
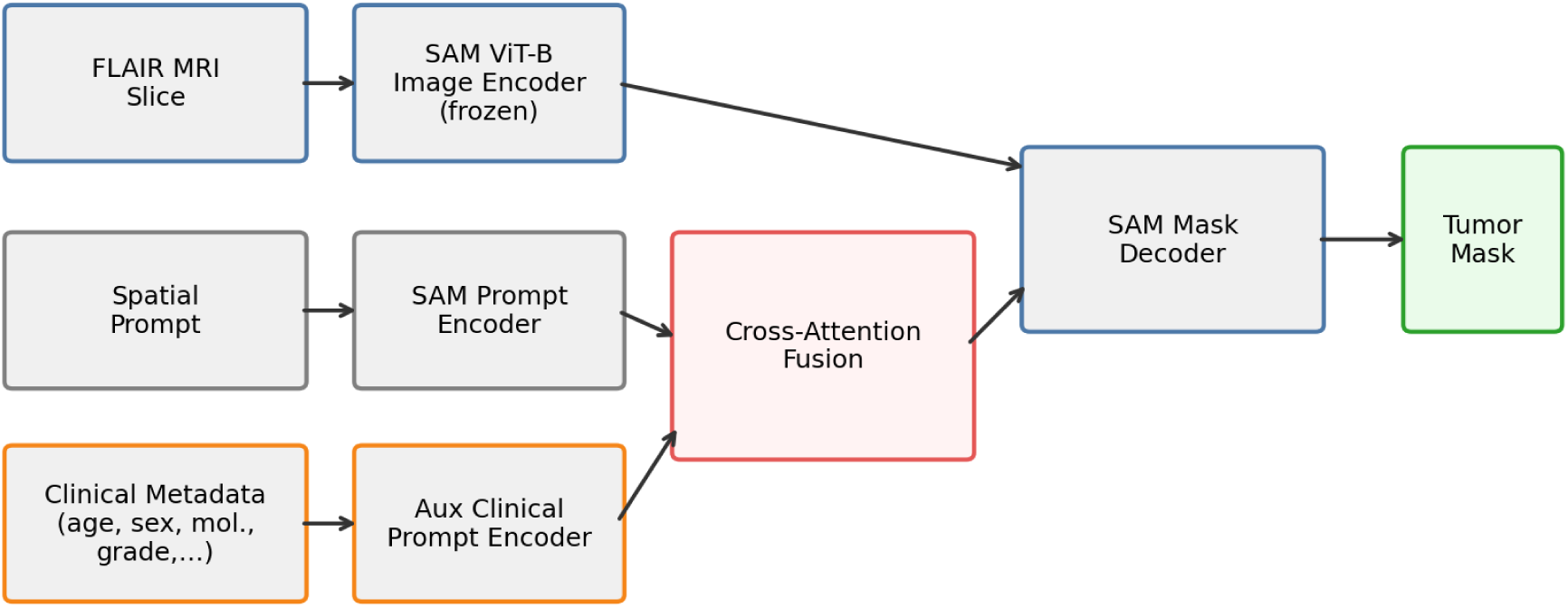
Framework overview: vision-language prompt-driven SAM framework with auxiliary clinical prompt integration.

**Fig. 2.**
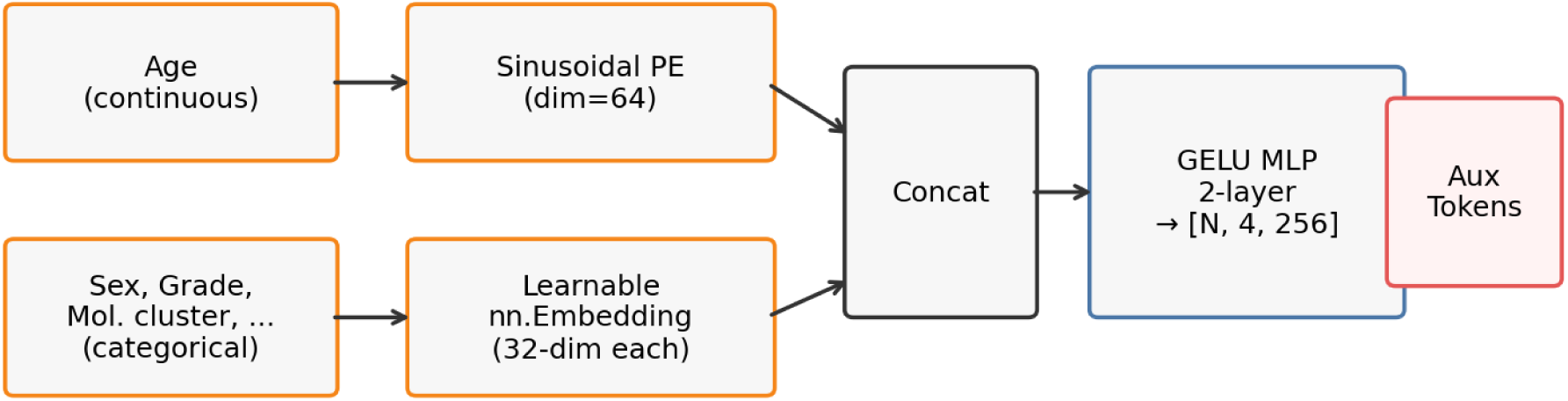
Detailed architecture of the Auxiliary Clinical Prompt Encoder.

**Fig. 3.**
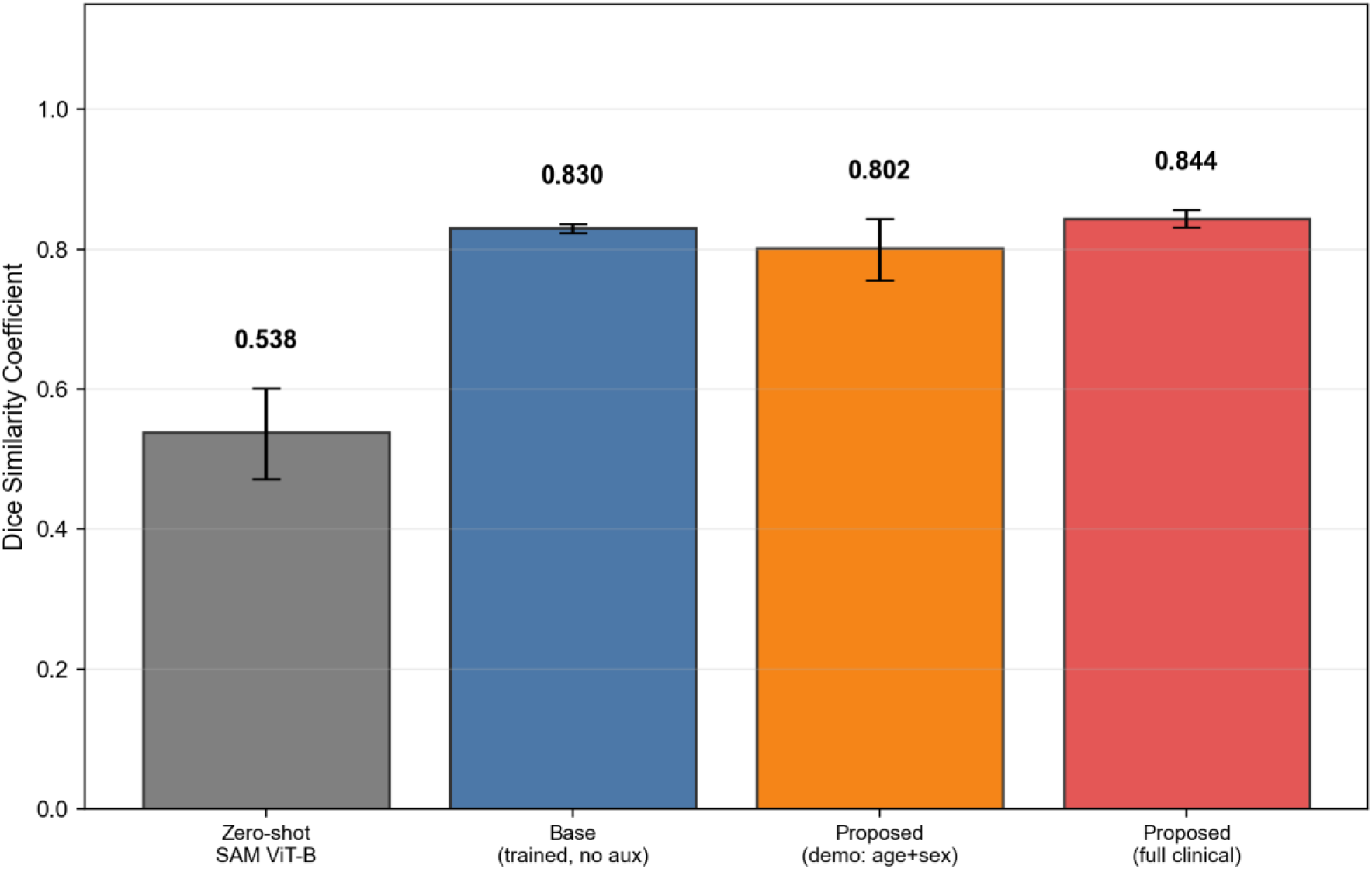
Bar chart comparing four configurations: zero-shot SAM, Base (trained without auxiliary information), Proposed (demographics only), and Proposed (full clinical). Base and Proposed (full) are evaluated under matched training and centroid-eval conditions from the robustness experiment (§3.4). Error bars indicate ± 1 SD across three seeds.

**Fig. 4.**
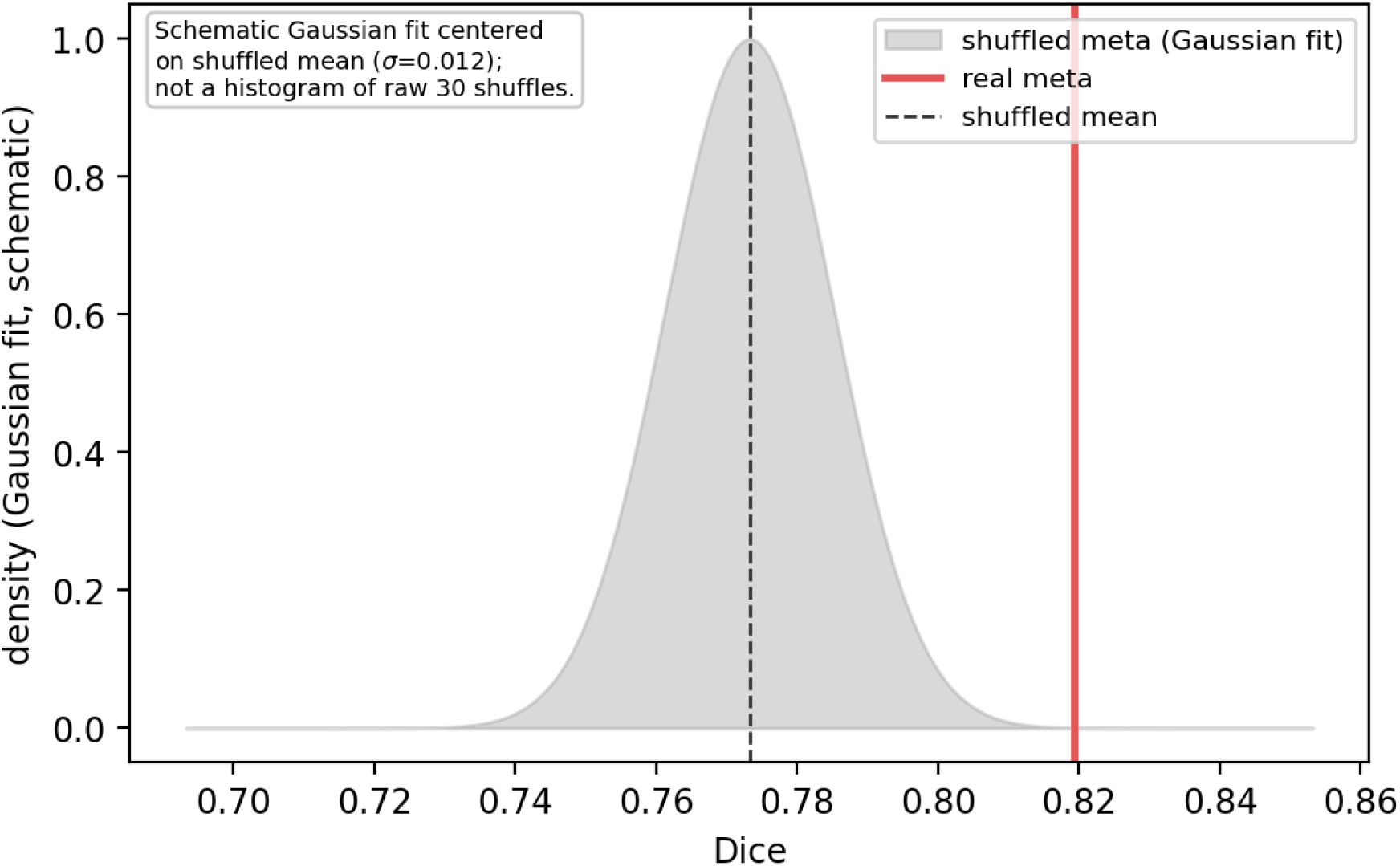
Permutation test on Proposed (full clinical). The schematic depicts a Gaussian fit centered on the shuffled mean for visualization purposes.

**Table 3.** Permutation test on Proposed (full clinical), seed 2025. Note: K = 30 shuffling trials; the empirical p-value 0.032 = 1/(K + 1) corresponds to the resolution limit of this test (see §2.6 and §4.5 (iv)).

| Condition | Dice |
| --- | --- |
| Real metadata (seed 2025) | 0.819 |
| Shuffled metadata (mean of $K=30$) | 0.773 |
| Empirical $p = (r+1)/(K+1)$ | 0.032 |

These results indicate that the trained Proposed (full clinical) model behaves differently under real metadata versus shuffled metadata, providing **preliminary evidence** that the auxiliary clinical prompts are effectively incorporated into the model’s predictions. However, with K = 30, the reported empirical p-value sits exactly at the lower-bound resolution of this test; the apparent nominal significance therefore reflects the maximum resolution of the test rather than a robust rejection of the null. Accordingly, this result is interpreted as **preliminary rather than conclusive** and should be re-validated with a substantially larger number of shuffles (K ≥ 1,000 is generally recommended [Phipson & Smyth, 2010]; see §4.4).

### 3.4 Robustness to spatial prompt degradation

To evaluate whether auxiliary clinical information can compensate for degraded or absent spatial prompts, we retrained both Base (no auxiliary information) and Proposed (full clinical) under a matched random-foreground-point training protocol (i.e., during training the point prompt was sampled uniformly from the foreground region, eliminating the train–eval mismatch that arises when training with random points but evaluating with the centroid) and evaluated them under six prompt-noise conditions: centroid (perfect), Gaussian noise with σ = 5, 10, and 20 pixels, random foreground point, and **no spatial prompt** (a dummy zero-coordinate point, forcing the model to rely entirely on image features and, in the case of Proposed, auxiliary clinical embeddings). Both models were evaluated under identical prompt conditions to ensure a fair comparison.

The results (Fig. 5) reveal two key findings. First, **Proposed (full clinical) outperformed Base under all six conditions** (Δ Dice ranging from +0.005 to +0.231), including the perfect-centroid condition (Δ = +0.014, 3/3 seeds). Second, and most strikingly, when the spatial prompt was completely removed, Base performance collapsed to Dice = 0.309 ± 0.019, whereas Proposed maintained Dice = 0.540 ± 0.051 — a difference of **Δ = +0.231 (seed-level paired-t p = 0.039, 3/3 seeds favoring Proposed)**. This indicates that the auxiliary clinical information provides the model with prior knowledge about expected tumor characteristics that can partially substitute for spatial guidance when the latter is unavailable.

**Fig. 5.**
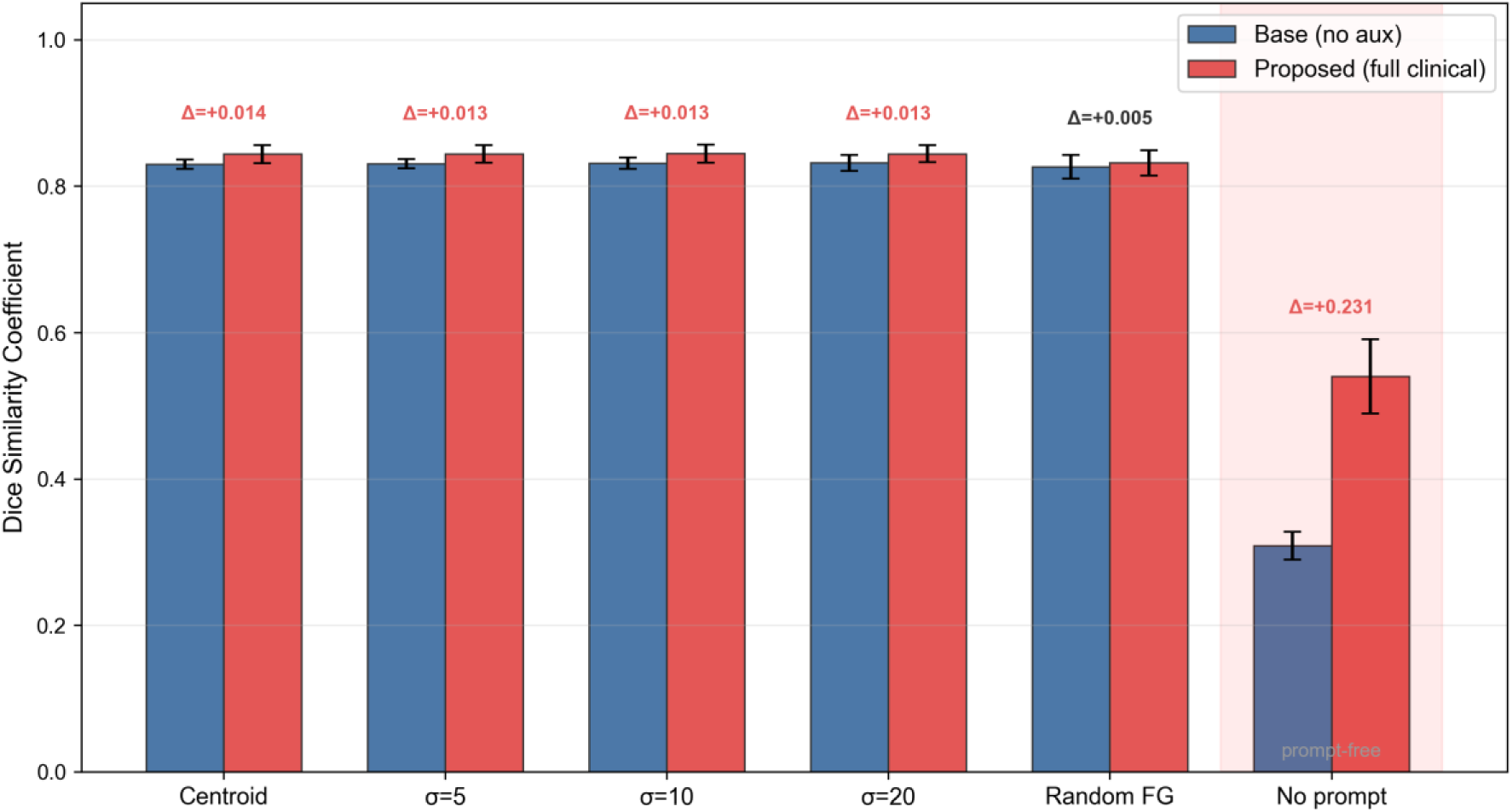
Robustness of Base vs Proposed (full clinical) across six spatial-prompt conditions. Error bars indicate ± 1 SD across three seeds.

These findings have important practical implications: in fully automated pipelines where no human operator provides a spatial click, the proposed framework can still produce meaningful segmentations by leveraging routinely available clinical metadata, whereas the base model without auxiliary information essentially fails. Under standard prompted conditions, the auxiliary information provides a consistent but modest improvement (Δ ≈ +0.01), suggesting that its primary value lies in robustness to prompt uncertainty rather than in peak-performance gains.

### 3.5 Per-sample behavior (secondary analysis)

To complement the aggregated results, per-sample behavior is visualized in Fig. 6 (paired Δ Dice) and Fig. 7 (Dice trajectory). Proposed (full clinical) maintained stable segmentation performance even in the low-Dice regime in which zero-shot SAM struggled, and the overall segmentation structure was consistently preserved when auxiliary clinical prompts were provided. These visualizations are intended as descriptive illustrations of model behavior; all statistical inference is based on the patient × seed level analyses of §3.1 and §3.4.

**Fig. 6.**
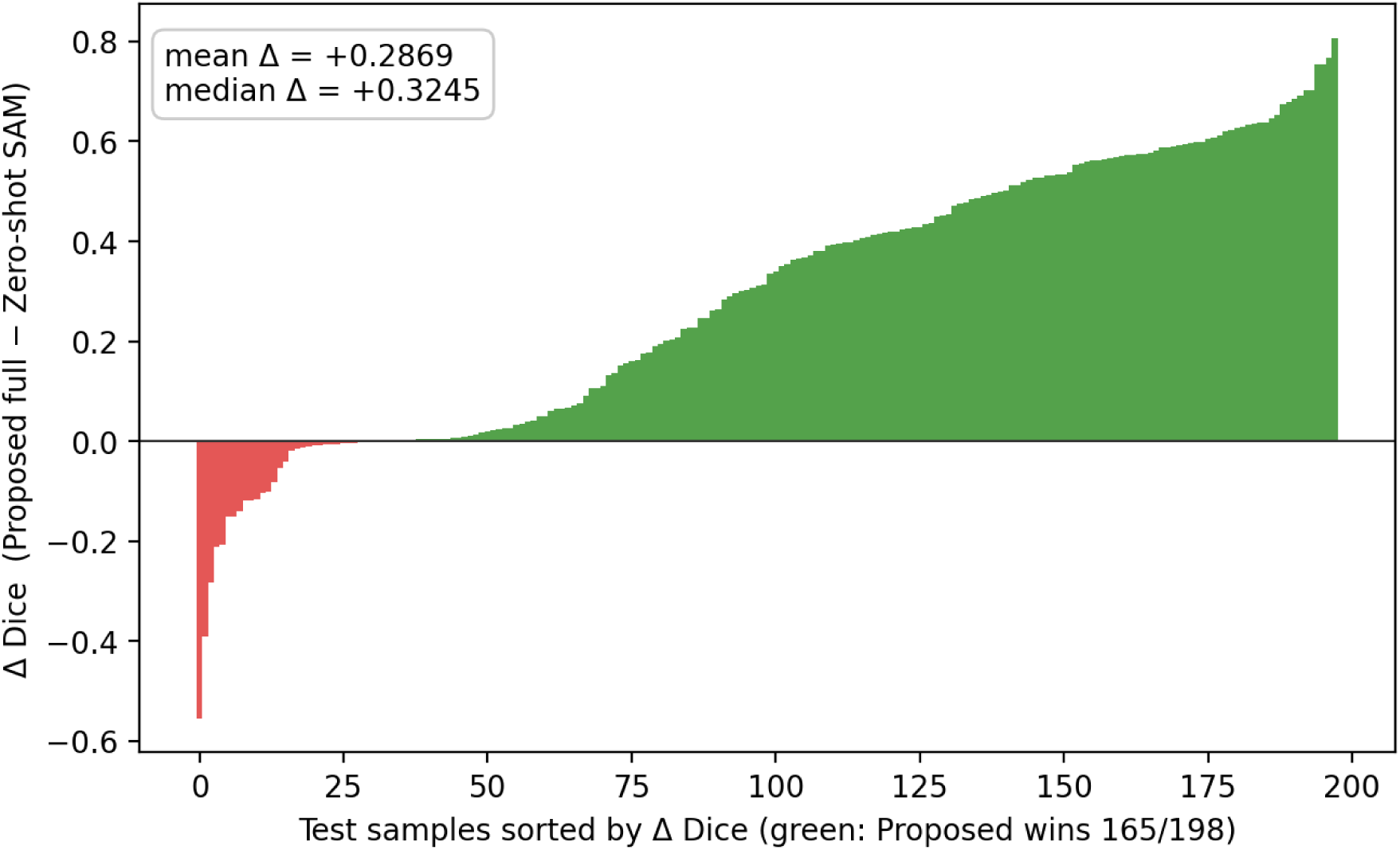
Per-sample paired Δ Dice (Proposed (full clinical) vs Zero-shot SAM, sorted).

**Fig. 7.**
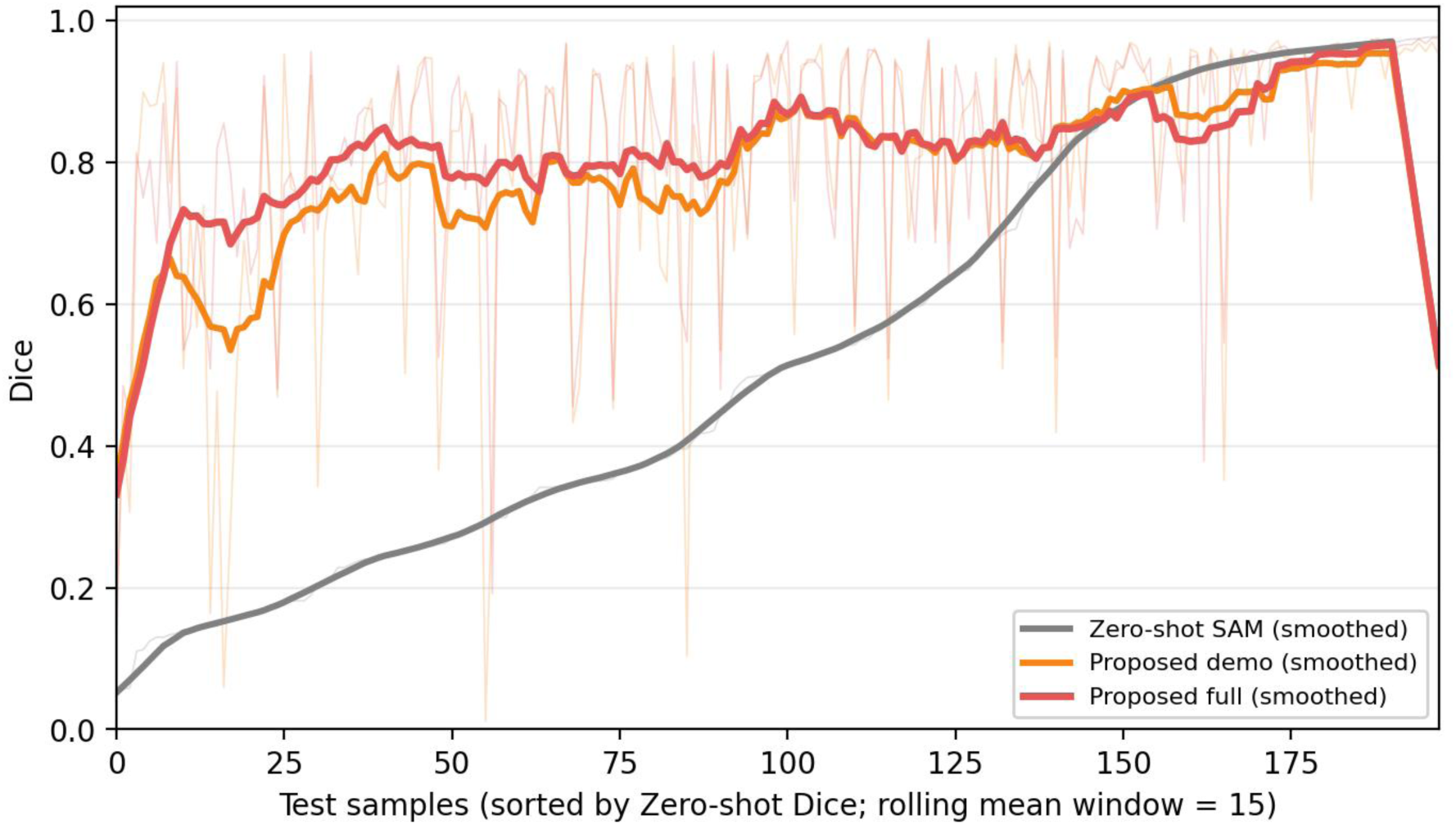
Per-sample Dice trajectories (rolling mean window = 15).

## 4. Discussion

### 4.1 Significance of auxiliary information integration

In this study, we showed that integrating auxiliary clinical information as multimodal prompts within a vision-language prompt-driven SAM extension framework substantially improves segmentation accuracy on the TCGA-LGG test set compared with the zero-shot SAM ViT-B baseline (Δ Dice = +0.287 at the patient × seed level). This study does not perform direct benchmark comparisons against established brain tumor segmentation models such as U-Net [8], nnU-Net [11], TransUNet [12], or Swin-UNet [13]; external benchmarking is left for future work (see §4.4). The present results nevertheless suggest that clinical data, which have rarely been considered as input in conventional segmentation pipelines, may contain information that contributes to estimating the imaging phenotype of tumors.

Approaches that rely solely on image information are known to reach a performance plateau in regions with unclear tumor boundaries or in highly infiltrative lesions. The fact that integrating auxiliary clinical information was associated with an improvement in accuracy in this study suggests that clinical data can capture biological properties of tumors that cannot be fully inferred from image cues alone; in other words, clinical data can serve the model as prior information originating from a non-image modality.

### 4.2 Verbalization and prompting of clinical reasoning

A notable strength of vision-language prompt-driven foundation models such as VLP-SAM is the flexibility with which diverse forms of information can be incorporated as natural-language input. In this study we integrated structured clinical data as prompts, and in principle this framework can be extended to ingest a wider range of information, although this extensibility has not been directly validated in the present experiments.

Particularly noteworthy is the possibility of leveraging, as a prompt, the clinical reasoning process that radiation oncologists employ when performing segmentation. In addition to imaging findings, radiation oncologists determine tumor boundaries by integrating knowledge of tumor extension patterns, anatomical relationships with adjacent structures, and prior treatment history. If such implicit reasoning processes could be explicitly verbalized and converted into prompts, the model could imitate expert decision strategies, which might yield further improvements in accuracy. Systematic investigation of methods for verbalizing clinical reasoning and of prompt design will therefore be required in future work.

### 4.3 Information selection and explainability

On the other hand, as the variety and quantity of integrated auxiliary information increase, the basis for the model’s decisions becomes increasingly opaque. The fact that it becomes unclear which input information contributes, and to what extent, to the segmentation output is a non-negligible issue from the viewpoint of reliability in clinical applications.

Ablation studies offer a useful approach for addressing this issue. The comparison between Proposed (full clinical) and Proposed (demographics) in this study did not reach statistical significance at the patient × seed level (§3.2), indicating that larger samples and more statistically powerful test plans are required to quantitatively separate the contributions of individual auxiliary categories. Furthermore, the permutation test of §3.3 provides preliminary evidence that the auxiliary information is actually utilized by the model, and it can be positioned as a complementary tool for evaluating the “utilization rate” of auxiliary information. Combining this complementary analysis with the conventional ablation approach allows both the necessity and the actual usage of auxiliary information to be assessed jointly.

In addition, combining these analyses with explainable AI (XAI) techniques will be important for visualizing which input information leads the model to classify a particular region as tumor, thereby presenting the decision rationale in a clinically interpretable form.

### 4.4 Clinical implications

If the positive signal observed in this study is confirmed in larger cohorts with external validation, the clinical implications are twofold. First, integrating routinely available clinical metadata into segmentation models could improve tumor delineation without requiring any additional data acquisition, thereby reducing the annotation burden on neuroradiologists. Second, the modular prompt-based design allows the system to be adapted to different clinical contexts (e.g., institutions with different molecular profiling panels) without retraining the image encoder, which may facilitate deployment across heterogeneous clinical environments. However, these implications remain speculative until direct comparisons with established clinical segmentation workflows and prospective evaluation studies have been conducted.

An important consideration is the **temporal availability of auxiliary information in clinical practice**. While demographics (age, sex) are available at the time of initial imaging, molecular profiling results (IDH mutation, methylation clusters) and definitive histopathological classification typically require tissue obtained through biopsy or resection — procedures that themselves rely on knowing the tumor location. This creates a potential chicken-and-egg issue for pre-operative use of the full clinical prompt. In practice, however, several deployment scenarios exist in which this concern is mitigated: (a) post-operative monitoring, where molecular and histological classification are already available from the initial surgery; (b) re-segmentation for radiotherapy planning after pathological confirmation; and (c) the demographics-only configuration (Proposed (demographics)), which uses only pre-operatively available information and already provides substantial improvement over the zero-shot baseline. Furthermore, regarding the risk of **information leakage**, the auxiliary clinical variables used in this study (age, sex, molecular cluster labels, histopathological grade) are patient-level metadata that do not directly encode spatial tumor boundary information; however, they are correlated with tumor morphology and location, and we cannot fully exclude the possibility that the model leverages these correlations as a shortcut rather than learning genuinely complementary features. Future work should include prompt attention visualization or gradient-based analyses to assess whether the model attends to prompt embeddings in a semantically meaningful way.

Regarding the non-significant ablation result (§3.2), we note that the current study may be underpowered (type II error risk) for detecting a small additional effect (Δ ≈ 0.023) of molecular and histopathological information over demographics alone. A post-hoc power analysis at d_z = 0.24 and α = 0.05 (two-sided) suggests that approximately 140 patient-seed observations would be required to achieve 80% power; our n = 66 provides only approximately 40% power for this comparison. The question of whether molecular information adds value beyond demographics therefore remains open and requires adequately powered studies.

### 4.5 Limitations

This study has the following important limitations.

**(i) Pseudo-replication and the unit of statistical analysis.** Initial analyses were based on paired t-tests over 198 per-slice observations (3 seeds × 22 patients × 3 slices), but these observations are not independent: slices from the same patient and observations from the same patient across different seeds are strongly correlated. In the current version, the primary analyses were re-run after aggregating the slice-level Dice and HD95 to the (seed, patient) unit (n = 66), and secondary analyses at the unique-patient level (n = 22) are also reported. The main claim (Δ Dice ≈ +0.287 for Proposed (full clinical) over zero-shot SAM) is robust across aggregation levels, whereas the smaller difference between Proposed (full clinical) and Proposed (demographics) (Δ ≈ +0.023) does not reach the α = 0.05 significance threshold at the patient × seed level (§3.2). A linear mixed-effects model with random effects for patient and seed would be an appropriate additional analysis in future work.
**(ii) Absence of direct comparison with external benchmarks.** The comparisons in this study are restricted to three configurations within our own framework (zero-shot SAM, Proposed (demographics), and Proposed (full clinical)). We did not benchmark directly against established brain tumor segmentation models such as U-Net [8], 3D U-Net [9], Attention U-Net [10], nnU-Net [11], TransUNet [12], or Swin-UNet [13]. Accordingly, we do not claim superiority over all existing methods; our findings should be interpreted strictly as an evaluation of the effect of integrating auxiliary clinical information into a vision-language prompt-driven foundation model.
**(iii) Domain gap in the zero-shot SAM baseline.** Zero-shot SAM ViT-B is a general-purpose model pre-trained on natural images, and its poor zero-shot performance on medical FLAIR MRI is consistent with previous reports [16, 17]. The HD95 of 218.2 px and Dice of 0.538 observed here therefore compare an unadapted foundation model against a medically trained model with auxiliary clinical information, rather than a strong medical baseline. Part of this apparent improvement may reflect the medical-domain adaptation of the base framework rather than the addition of auxiliary information per se.
**(iv) Underpowered permutation test (K = 30).** The permutation test presented in §3.3 uses only K = 30 shuffles and therefore has a resolution-limited lower bound on its empirical p-value of (r + 1) / (K + 1) = 1/31 ≈ 0.032. The reported p = 0.032 sits exactly at this limit. For a rigorous significance test we recommend re-running the test with K ≥ 1,000 [Phipson & Smyth, 2010]. The current result should be regarded as **preliminary evidence** of effective use of auxiliary information.
**(v) Dataset scope and diversity.** The evaluation is limited to TCGA-LGG (Buda Kaggle subset, n = 110 patients, WHO grade II–III), and validation on larger multi-center, multi-country cohorts is required. Our cohort does not contain glioblastoma (glioblastoma, IDH-wildtype, WHO CNS grade 4), so generalization to the prognostically most relevant adult diffuse glioma subtype requires separate validation.
**(vi) Single modality and single scanner source.** This study uses FLAIR alone; multi-sequence evaluation (T1, T1-contrast, T2, FLAIR) remains future work. Robustness to scanner and protocol heterogeneity has not been evaluated either.
**(vii) Surrogate nature of molecular information.** The molecular cluster variables used in this study (MethylationCluster, RNASeqCluster, miRNACluster) are unsupervised cluster assignments derived from TCGA high-throughput profiling and correspond to, but are not identical with, the direct molecular markers (IDH mutation, 1p/19q co-deletion, MGMT promoter methylation by direct assay, TERT promoter mutation) specified by the 2021 WHO classification [3]. Evaluation using direct molecular assays as auxiliary input is an important future task.
**(viii) Computational resources and backbone capacity.** The study operates under a single-GPU constraint (NVIDIA RTX 4060 Laptop, 8 GB VRAM) and adopts SAM ViT-B as the backbone. Re-evaluation with larger backbones (SAM ViT-L / ViT-H) and with medically pre-trained alternatives such as MedSAM [18] or SAM-Med2D [19] is left for future work.
**(ix) Retrospective nature.** This is a retrospective methodological study based on secondary analysis of existing public data; prospective validation in real clinical settings is a subsequent step.

### 4.6 Future perspectives

Building on the findings of this study, several directions can be considered for future work. First, direct benchmark comparisons with established brain tumor segmentation models (e.g., nnU-Net) will position the effect of auxiliary information integration against a stronger external reference. Second, rigorous re-analysis with K ≥ 1,000 permutations and with linear mixed-effects models should quantify the effective use of auxiliary information and the contributions of individual categories. Third, external validation on larger, multi-center, multi-country cohorts with multi-sequence MRI (T1/T1c/T2/FLAIR) that includes both LGG and GBM (IDH-wildtype, WHO CNS grade 4) is needed to evaluate generalizability. Fourth, the contribution of direct molecular assays (IDH, MGMT, TERT, etc.) as auxiliary inputs should be assessed.

In view of potential clinical deployment, ensuring explainability is an indispensable requirement. By combining model designs that output decision rationales with the insights from the permutation tests and ablation studies presented here, it may be possible to build a segmentation system that reconciles accuracy and interpretability. Such efforts may help the proposed framework contribute to the future development of clinical decision-support tools in the diagnosis and treatment planning of brain tumors, although practical deployment would require multi-center prospective validation and regulatory review that are beyond the scope of the present study.

## 5. Conclusions

In this study, building on a vision-language prompt-driven SAM extension framework, we proposed a lower-grade glioma MRI segmentation method that integrates patient demographics, TCGA-derived molecular cluster variables, and histopathological parameters as multimodal prompts, and we evaluated it on the TCGA-LGG dataset. The primary analysis at the patient × seed level (n = 66) showed that Proposed (full clinical) improved over the zero-shot SAM ViT-B baseline by Δ Dice = +0.287 (Bonferroni-corrected p ≪ 0.001, Cohen’s d_z = +1.25, Wilcoxon p = 2.0 × 10⁻¹⁰), robust under unique-patient aggregation (n = 22) and linear mixed-effects modeling (LME p = 5.7 × 10⁻²⁰). The additional contribution of the full clinical configuration over demographics alone (Δ Dice = +0.023) remained a directional trend that did not reach statistical significance in this cohort.

Crucially, a robustness analysis demonstrated that Proposed (full clinical) **outperformed the trained Base model (no auxiliary information) under all six tested spatial-prompt conditions**, with the advantage being most pronounced in the prompt-free regime: when the spatial point prompt was entirely removed, Base collapsed to Dice = 0.309 while Proposed maintained Dice = 0.540 (Δ = +0.231, seed-level paired-t p = 0.039, 3/3 seeds). This finding indicates that auxiliary clinical information can partially substitute for spatial guidance by providing the model with prior knowledge about expected tumor characteristics, enabling meaningful segmentation even in fully automated pipelines without human interaction.

Direct benchmarking against established brain tumor segmentation models, external validation on multi-center multi-modality cohorts, and permutation tests with a substantially larger K are left for future work. The prompt-driven architecture is readily extensible to additional input types, and, pending external validation, the framework may contribute to the future development of clinical decision-support tools for the diagnosis and treatment planning of brain tumors.

## Declarations

### Ethics statement (IRB / Institutional review)

This study involves only **secondary analysis of an existing, publicly available, fully anonymized dataset** and does not include any recruitment of new patients, new data collection, intervention, acquisition of biological specimens, or access to identifiable personal health information (PHI).

The medical imaging data used in this study are from The Cancer Genome Atlas (TCGA), accessed through The Cancer Imaging Archive (TCIA) [23] and through the TCGA-LGG MRI Segmentation subset [24] released on Kaggle by Buda et al. These data were originally collected under protocols approved by the **Institutional Review Board (IRB) or equivalent independent ethics committee** of each contributing institution, and written informed consent was obtained from all participants at the time of data collection. De-identification was completed by the original data providers.

This study falls under the exemption provision of the U.S. Department of Health and Human Services (HHS) **Common Rule (45 CFR 46.104(d)(4))** concerning “research involving the collection or study of existing data, documents, records (…) if these sources are publicly available or if the information is recorded by the investigator in such a manner that subjects cannot be identified, directly or through identifiers linked to the subjects,” and therefore no additional IRB approval, ethics review, or participant consent was required. The design and conduct of this study adhered to the Declaration of Helsinki (2013 revision) and to all applicable institutional policies and international guidelines.

In accordance with the ethics policies of the authors’ affiliated institution, this study has been confirmed to qualify for IRB-exempt status.

### Clinical trial registration statement

This study is **not an interventional study and does not constitute a clinical trial**. It is a retrospective methodological study (development and evaluation of a deep learning model) using an existing, publicly available, anonymized dataset, and does not involve recruitment of new subjects, any intervention, or any follow-up. Therefore, under the ICMJE definition (https://www.icmje.org/recommendations/browse/publishing-and-editorial-issues/clinical-trial-registration.html), **clinical trial registration is not required** and no trial registration ID has been assigned.

### Patient identifiability and de-identification statement

All medical imaging data used in this study originate from a **fully anonymized public dataset** (TCGA-LGG via the Kaggle release by Buda et al. [24]).

- The data used do not contain patient names, dates of birth, addresses, medical record numbers, social security numbers, or any of the other HIPAA Safe Harbor 18 identifiers.
- Image metadata such as DICOM headers were de-identified in advance by the data provider, and this study does not handle any additional patient-identifying metadata.
- Patient age is included in the original dataset as an integer value (age_at_initial_pathologic), but no case in this cohort falls under the HIPAA “ages over 89” rule.
- All files released as part of the results of this study (the main text, figures and tables, supplementary materials, source code, and trained model weights) contain no personally identifiable information.
- Dataset-specific patient identifiers (TCGA Patient IDs) are not individually disclosed in this manuscript; only aggregated statistics and per-sample identifiers in the format TCGA_* s{N} are reported in the paper and supplementary materials. These identifiers were already released as de-identified IDs at the time of TCIA publication.

The authors confirm that this manuscript and its accompanying materials **do not create any risk of direct or indirect re-identification of patients**.

### Patient and public involvement (PPI) statement

This study is an analysis of a publicly available, anonymized secondary dataset, and no direct Patient and Public Involvement (PPI) took place at any stage of study design, conduct, analysis, or reporting. Because this study does not involve recruitment of new patients, PPI activities are not applicable.

### Consent for publication

This study does not contain any new patient data, new images, identifiable personal information, or case descriptions, and therefore individual patient consent for publication is not required. Broad research-use consent obtained at the time of release of the original dataset covers this use.

### Distribution license

This preprint is distributed under the **Creative Commons Attribution 4.0 International License (CC BY 4.0)**. Users are permitted to copy, modify, redistribute, and reuse the manuscript in any form, including for commercial purposes, provided that the original work is properly cited.

### Data availability statement

The TCGA brain tumor imaging data used in this study are publicly available through The Cancer Imaging Archive (TCIA) (https://www.cancerimagingarchive.net/). We used the TCGA-LGG MRI Segmentation subset released on Kaggle by Buda et al. (https://www.kaggle.com/datasets/mateuszbuda/lgg-mri-segmentation). The TCGA-LGG collection is accessible for research purposes without restriction. The source code, trained model weights, and experimental logs of this study will be released upon acceptance through GitHub and a dedicated model repository (https://github.com/chordix/vlp-sam-aux-clinical). In the interim, materials necessary for reproduction are available from the corresponding authors upon reasonable request.

### Code availability statement

All implementations used in this study were written in Python 3.11 with PyTorch 2.3.1 and CUDA 12.1, and will be released in the aforementioned GitHub repository upon acceptance.

### Funding statement

This research received no specific grant from any funding agency in the public, commercial, or not-for-profit sectors. The work was conducted as part of the in-house research and development activities of the authors’ affiliation, **Chordix Inc.**. The institution had no independent role in the study design, data analysis, interpretation of results, manuscript writing, or the decision to submit the manuscript for publication; the research was performed and reported independently in accordance with scientific integrity.

### Competing interests statement

The authors are affiliated with **Chordix Inc.**, a medical AI startup that develops clinical decision-support technologies. This study was conducted as part of the company’s in-house technology development program. Although this research may inform the development of future commercial products, the study design, data analysis, and reporting of results were conducted independently of commercial considerations. The authors declare that they have not received, in the past 36 months, any payments or services from any third party that could influence, or be perceived as influencing, the submitted work (None to declare).

### Author contributions (CRediT)

**Yuto Hakata**: Conceptualization, Methodology, Software, Investigation, Formal analysis, Writing – original draft, Visualization

**Miko Oikawa**: Methodology, Software, Validation, Investigation, Formal analysis, Writing – original draft

**Shin Fujisawa**: Review & editing

## Data Availability

The TCGA-LGG MRI Segmentation dataset is publicly available on Kaggle (https://www.kaggle.com/datasets/mateuszbuda/lgg-mri-segmentation). All code and trained model weights will be made available upon reasonable request to the corresponding authors.

https://www.kaggle.com/datasets/mateuszbuda/lgg-mri-segmentation

## Acknowledgments

We thank Buda et al. for releasing the TCGA-LGG MRI Segmentation dataset used in this study, as well as the operational team of The Cancer Imaging Archive (TCIA).

## Notes

### Competing Interest Statement

The authors have declared no competing interest.

### Funding Statement

This study did not receive any external funding. The work was conducted as in-house research at Chordix Inc. No payments or services were received from any third party for any aspect of this work.

### Author Declarations

This study used only openly available human data. The TCGA-LGG MRI Segmentation dataset was publicly released on Kaggle by Buda et al. prior to the initiation of this study and can be downloaded at: https://www.kaggle.com/datasets/mateuszbuda/lgg-mri-segmentation. The original TCGA-LGG clinical and genomic data are available from the Genomic Data Commons (GDC) portal at: https://portal.gdc.cancer.gov/projects/TCGA-LGG.

